# Vascular brain injury modifies the relationship between sleep duration, cognition, and white matter hyperintensity burden in the Alzheimer’s disease continuum

**DOI:** 10.64898/2026.03.12.26348239

**Authors:** Soraya Lahlou, Zahinoor Ismail, Eric E. Smith, Thien Thanh Dang-Vu, AmanPreet Badhwar

**Affiliations:** Multiomics Investigation of Neurodegenerative Diseases (MIND) Laboratory, Montréal, QC, Canada; Centre de Recherche de l’Institut Universitaire de Gériatrie de Montréal (CRIUGM), Montréal, QC, Canada; Department of Psychiatry, Hotchkiss Brain Institute, University of Calgary, Calgary, AB, Canada; Department of Clinical Neurosciences, Hotchkiss Brain Institute, University of Calgary, Calgary, AB, Canada; Sleep, Cognition and Neuroimaging Laboratory, Department of Health, Kinesiology and Applied Physiology, Concordia University, Montréal, QC; Département de pharmacologie et physiologie, Faculté de médecine, Université de Montréal, Montréal, Canada; Institut de génie biomédical, Université de Montréal, Montréal, Canada

**Keywords:** alzheimer’s disease, vascular brain injury, white matter hyperintensities, sleep duration, cognition

## Abstract

**INTRODUCTION:** Self-reported sleep duration is associated with dementia-risk and vascular brain injury markers, including white matter hyperintensities (WMHs), yet how additional cerebrovascular pathology alters these relationships remains unclear.

**METHOD:** In a deeply-phenotyped cohort (735) including healthy and Alzheimer’s continuum individuals (subjective and mild cognitive impairment (MCI), dementia (AD), MCI and AD with high vascular burden (+V)), sleep was assessed using the Pittsburgh Sleep Quality Index. Individuals were classified as WMH-only or WMH+ (WMHs with microbleeds, infarcts, or cerebral amyloid angiopathy). Linear models tested interactions between sleep duration, WMH+ status, and diagnostic group on WMH burden and cognition.

**RESULTS:** In MCI+V and AD, WMH+ significantly amplified the association between shorter sleep and greater WMH burden. In AD, longer sleep related to better cognition in WMH+, but worse cognition in WMH-only (exploratory).

**DISCUSSION:** Additional vascular brain injury modifies how sleep relates to WMH burden and cognition across the AD continuum, highlighting the importance of moving beyond WMHs alone.

## 1 INTRODUCTION

Vascular brain injury is widespread in older adults and is increasingly recognized as an early contributor to multiple neurodegenerative diseases (NDDs)^1^, particularly Alzheimer’s disease (AD)^2^. On neuroimaging, it manifests as white matter hyperintensities (WMHs), lacunes, microbleeds, and infarcts - markers that are associated with increased dementia risk, including AD^3,4^. Although these markers may reflect distinct etiologies^5^ and risk profiles^6^, they frequently co-occur in AD^7,8^. Among them, WMHs appear early^9–11^ and have recently been validated as a core neuroimaging marker of vascular brain injury in AD^12^. Together, these markers provide critical insight into the vascular contribution to cognitive decline in AD, especially when considered alongside other factors associated with dementia risk, such as sleep.

Sleep disturbances, like vascular brain injury, are common in aging and NDDs, and have been associated with dementia^13^. Large cohort studies and meta-analyses demonstrate a U-shaped association between self-reported sleep duration and dementia, including AD, with both short and long sleep linked to cognitive decline^14–16^ — a pattern mirrored by cerebrospinal fluid (CSF) amyloid, an established biomarker of AD pathology^17^. These findings suggest that deviations in sleep duration may signal early AD-related processes, although it remains unclear whether this contributes to disease development or reflects downstream effects of underlying pathology.

While evidence supports the detrimental effects of sleep deprivation on cognition, through mechanisms such as accumulation of hallmark AD proteins (amyloid, tau) and inflammation^18,19^, the biological basis linking long sleep to cognitive decline is less understood. One prevailing hypothesis is that longer sleep reflects a compensatory response to underlying AD processes^20,21^.

Supporting this, in individuals with a parental history of AD, longer sleep has been associated with elevated CSF markers of AD pathology, yet better cognitive performance, suggesting compensatory processes may be at play^20^.

Given the strong links between sleep duration and AD risk, recent work has begun to examine how sleep duration relates to markers of vascular brain injury, particularly WMHs, in cognitively unimpaired (CU) adults. To date, four large UK BioBank sampling of dementia-free adults have shown a U-shaped relationship between self-reported sleep duration and WMH burden^22–25^. Longitudinally, Baril and colleagues^26^ demonstrated that increases in sleep duration predicted future WMH burden — but only when moving from normal to longer sleep. Furthermore, the association between longer sleep and WMH burden is amplified in individuals with vascular risk factors (e.g., hypertension^25^, diabetes^27^), suggesting a stronger link between longer sleep and vascular brain injury in those with existing vascular risk. While these studies provide insight on the links between sleep and WMHs in CU^22–29^, it remains unclear how the relationship between sleep, WMHs and cognition evolve across the AD continuum.

Emerging literature also suggests that the relationship between longer sleep and vascular brain injury may not be uniform, and may depend on the type or overall burden of injury. In individuals following mild stroke, longer sleep was associated with greater microbleed burden, but not WMHs, and predicted worse cognitive outcomes^30^. This absence of association with WMHs suggests that considering only WMHs may not be sufficient, and that longer sleep may not always reflect compensation. This highlights an important question: beyond WMHs, is the presence of additional vascular brain injury associated with sleep, and does it modify the relationship between sleep, WMH burden and cognition?

Based on prior evidence that longer sleep may reflect underlying disease and its link to vascular brain injury, we tested three hypotheses across the AD continuum. First, sleep duration is longer in individuals with additional vascular brain injury beyond WMHs (WMH+). Second, WMH+ amplifies the relationship between longer sleep and higher WMH burden. Third, WMH+ will also amplify the relationship between longer sleep and worse cognition (exploratory).

To test these hypotheses we leveraged a deeply phenotyped pan-Canadian cohort spanning the AD continuum. Given sleep is a modifiable factor^31,32^, understanding how vascular brain injury shapes these associations may help inform its use as a potential therapeutic target.

## 2 METHOD

### 2.1 Cohort

Using the Canadian Consortium for Neurodegeneration in Aging (CCNA)-COMPASS-ND cohort^33^, we analyzed individuals across the AD continuum without and with high vascular brain injury burden (+V). This cohort is well-suited to examine the cerebrovascular contributions to sleep and cognition given its detailed brain vascular assessments and cognitive phenotyping^34^. All included participants had automatically segmented WMH data, radiological assessment of vascular brain injury, self-reported sleep measures, and a comprehensive neuropsychological evaluation^35^, with two-year follow-up.

The sample comprised six diagnostic groups: CU (N=105, 77% F), subjective cognitive impairment (SCI; N=133, 73% F), mild cognitive impairment (MCI; N=234, 40% F), mild cognitive impairment with high vascular burden (MCI+V; N=131, 44% F), AD (N=90, 42% F) and AD with high vascular burden (AD+V; N=43, 44% F). One participant in the AD group was excluded due to an implausible sleep duration value.

Diagnostic criteria for CU and SCI were defined by unimpaired cognitive performance, indicated by a Global Clinical Dementia Rating (CDR) score of 0, a Montreal Cognitive Assessment (MoCA) score above 25, a CERAD score above 5, and performance on the Logical Memory II above that of the ADNI education-adjusted cutoffs (for details, see paper on the COMPASS-ND neuropsychological battery^35^). Additionally, SCI was defined as the presence of subjective cognitive complaints accompanied with concern about these changes, as per the Jessen criteria^36^. MCI was defined as impaired cognition on at least one cognitive test (Global CDR > 0, MoCA 13-24, CERAD <6, or Logical Memory II below ADNI education-adjusted cutoffs). AD was defined by impairment on at least two of the above cognitive tests, in addition to a positive response to the question “Does the participant have cognitive deficits that interfere with independence in everyday activities such as paying bills or managing medications?”. Participants received a ‘+V’ designation if they were identified as having a significant vascular component to their cognitive impairment (i.e., strategic infarcts, two or more supratentorial brain infarcts or confluent WMHs)^34^.

### 2.2 Sleep assessment

Self-reported sleep was measured using the Pittsburgh Sleep Quality Index (PSQI) questionnaire^37^ at both timepoints. The PSQI is a self-reported questionnaire which assesses overall sleep characteristics during the last 4 weeks across seven sub-categories: sleep quality, sleep latency, sleep duration, sleep efficiency, sleep disturbances, use of sleeping medications and daytime dysfunction. The PSQI is intended as a screening tool, to help discriminate between good and poor sleepers. In addition to the PSQI, participants also reported the presence of a sleep breathing disorder as part of their medical history assessment.

### 2.3 Neuroimaging of vascular brain injury

The CCNA-COMPASS-ND cohort is scanned following the Canadian Dementia Imaging Protocol^38,39^, designed to reduce inter-site variability in multi-centre studies, using 3T scanners (GE, Philips, Siemens).

#### 2.3.1 MRI preprocessing, WMH segmentation, and volumetric quantification

Sequences used to detect WMH included 3D isotropic T1-weighted and 2D T2-weighted fluid attenuated inversion recovery (FLAIR). Images were denoised^40^, corrected for intensity non-uniformity^41^ and intensity-normalized, as described previously^42^. FLAIR and T1-weighted images were co-registered using a 6-parameter rigid-body registration and mutual information cost function^42^. Images were visually inspected for artefacts. WMHs were segmented using a publicly available automated pipeline^43,44^ in native FLAIR space, with visual quality control of segmentation masks and subsequent volume calculation.

#### 2.3.2 MRI-based radiological assessment of vascular brain injury and composite burden derivation

We leveraged available radiological reports^34^ detailing the presence of infarcts, microbleeds and/or fulfillment of the Boston criteria for possible or probable cerebral amyloid angiopathy (CAA) ^45,46^ to derive a composite measure of overall vascular brain injury burden. Each feature was coded as present or absent, yielding a maximum composite burden score of 3. For interpretability, additional vascular brain injury burden among individuals with WMHs was binarized as WMH-only (WMHs without additional vascular brain injury) or WMH+ (WMHs with additional vascular brain injury). This WMH-only vs WMH+ classification complements the clinical +V designation described above in Section 2.1.

### 2.4 Statistics

Analyses were performed using RStudio^47^ (version 4.0.5). Total WMH volumes were normalized using log-transformation and are referred to as WMH burden. We examined the relationship between sleep measures derived from the PSQI questionnaire and WMH burden. As most prior studies report findings using self-reported sleep duration, this variable was prioritized for primary analyses, with other PSQI domains kept as exploratory analyses. We provide a schematic of the analyses performed in this paper in Figure 1.

**Figure 1.**
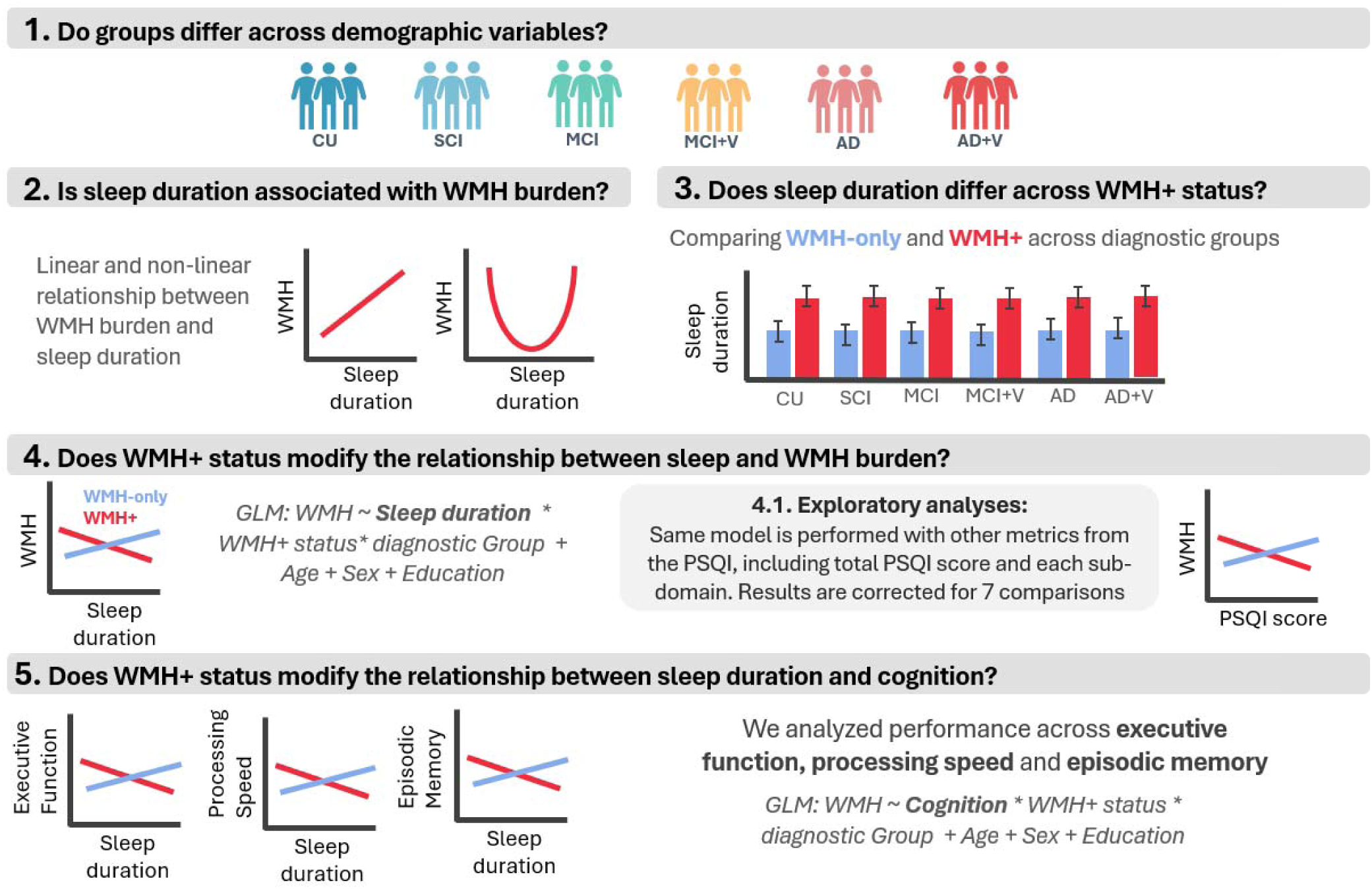
Schematic of analyses. (1) Demographic across groups (2) Linear and non-linear relationship between sleep duration and WMH burden. (3) Comparison of sleep duration across WMH+ status within each diagnostic group. (4) Moderating effect of WMH+ status on th relationship between sleep duration and WMH burden, with exploratory analyses using PSQI score and subdomains. (5) Moderating effect of WMH+ status on the relationship between sleep duration and cognitive performance at baseline and at follow-up.

#### 2.4.1 Demographics

Descriptive statistics are summarized for demographic, clinical and neuropsychological characteristics across our six diagnostic groups. Group differences were assessed with Kruskal-Wallis tests for continuous variables, and Chi-Square tests for categorical variables. When significant, post-hoc pairwise comparisons were performed using either Wilcoxon rank test or proportion test, for continuous and categorical variables respectively, followed by FDR correction.

#### 2.4.2. Relationship between sleep duration and WMH burden

We first modeled the non-linear relationship between self-reported sleep duration and WMH burden within each diagnostic group to replicate the previously reported U-shaped association. Restricted cubic spline models were fitted using the R packages “rms” and “rmsMD”, with three knots selected to minimize overfitting in smaller samples^48^. Models were adjusted for age, sex, and education. We then repeated the analysis across all participants, irrespective of the diagnostic group. Model fit was evaluated by comparing the spline model to a linear model (including sleep duration as a linear term) using ANOVA.

#### 2.4.3. Difference in sleep duration across WMH+ status

To characterize the relationship between sleep duration and vascular brain injury beyond WMH burden, we examined whether participants classified as WMH-only or WMH+ showed differences in sleep duration using ANOVAs controlling for age, sex, and education.

#### 2.4.4. Moderating effect of WMH+ status on the relationship between sleep duration and WMH burden

To test whether WMH+ status modified the relationship between sleep duration and WMH burden, we ran a linear regression including WMH+ as an interaction term with sleep duration and diagnostic group (WMH burden ∼ sleep duration * WMH+ status * diagnostic group + Age + Sex + Education). Given our goal was to test the modifying effects of WMH+, the interaction terms were our primary focus. Diagnostic group was dummy-coded with CU participants as the reference, and sex was coded as a factor with “Female” as the reference. Given that sleep breathing disorder is a cerebrovascular risk factor^49^, we conducted a sensitivity analysis including it as a covariate. To capture the full spectrum of vascular brain injury, we additionally report results from a more complex model treating the composite burden score (based on infarcts, microbleeds, and CAA) as a four-level factor (scores 0–3) in the Supplementary Material.

#### 2.4.4.1 Moderating effect of WMH+ status on the relationship between PSQI subdomains and WMH burden

We also investigated interactions between WMH+ status and other self-report measures of sleep, besides sleep duration. Seven separate linear models were fit for total PSQI score and each of six PSQI sub-domains (sleep quality, latency, efficiency, disturbances, daytime disturbances, and sleep medication), excluding sleep duration to avoid redundancy. (e.g., WMH burden ∼ sleep variable * WMH+ status* diagnostic group + Age + Sex + Education). Given the main analyses focused on self-reported sleep duration, we excluded the PSQI sub-domain of sleep duration in these analyses. FDR correction was applied across these seven models.

#### 2.4.5. Moderating effect of WMH+ status on the relationship between sleep duration and cognitive performance

To examine whether WMH+ status modified the relationship between sleep duration and cognitive performance at baseline, we fit a separate linear regression testing the interaction between sleep duration and WMH+ status on cognition (Cognition ∼ sleep duration * WMH+ status * diagnostic group + Age + Sex + Education). Significant interactions were further explored using simple-slope analysis to characterize effect direction and magnitude. Cognitive analyses focused on tests assessing either processing speed, executive function or episodic memory, given evidence that these are among the first to decline in the AD continuum^50,51^. Namely, we used performance on the Trail Making Test (TMT) Parts A and B, the Choice Reaction Time (CRT) test, the Inhibition trials of the Stroop, and the immediate and delayed recall on the Rey Auditory Verbal Learning (RAVLT) test. Scores were scaled to age- or age-and education-specific norms^35^ and reverse-coded so that higher values reflect better performance. False discovery rate (FDR) correction was applied at 0.05 across six cognitive tests.

Finally, we tested whether baseline sleep duration and its interaction with WMH+ status predicted cognitive performance at follow-up (Follow-up cognition ∼ sleep duration * WMH+ status * diagnostic group + Age + Sex + Education), and diagnostic conversion among CU or SCI participants at baseline.

## 3 RESULTS

### 3.1 Cohort demographics

Demographic information is summarized in Table 1 across the six diagnostic groups. At baseline, participants’ age ranged from 69.9 to 78.9 years and whole-brain WMH volume ranged from 8.39 cm^3^ to 32.5 cm^3^, with both age and WMH burden increasing across the AD continuum (p>0.00001). Self-reported sleep duration also showed a significant increase from the CU to the AD groups (p>0.00001), ranging from 6.94 hours in the CU group to 8.27 hours in the AD+V group (Supplementary Figure 1).

**Table 1.** Cohort demographics. ^a^ Kruskal-Wallis test for continuous variables. When results were significant, post-hoc pairwise Wilcoxon rank tests with FDR correction were performed. ^b^ Chi-Square test for categorical variables. When results were significant, post-hoc pairwise proportion tests with FDR correction were performed.

|  | CU<br>(N=105) | SCI<br>(N=133) | MCI<br>(N=234) | MCI+V<br>(N=131) | AD<br>(N=89) | AD+V<br>(N=43) | p-value | Post-hoc pairwise<br>comparisons |
| --- | --- | --- | --- | --- | --- | --- | --- | --- |
| Age (SD) <sup>a</sup> | 69.6 (6.09) | 70.2 (6.01) | 72.0 (6.72) | 76.3 (6.31) | 72.8 (7.76) | 78.9 (5.63) | >0.00001 | CU, SCI < MCI, AD < MCI+V < AD+V |
| Sex, % Female (N) <sup>b</sup> | 77% (81) | 73% (97) | 40% (94) | 44% (58) | 43%<br>(N=38) | 44% (19) | >0.00001 | AD, AD+V < MCI, MCI+V < CU, SCI |
| Education (SD) <sup>a</sup> | 15.3 (2.28) | 15.8 (2.51) | 14.9 (2.98) | 14.8 (3.03) | 14.9 (2.92) | 15.0 (2.74) | 0.07 | - |
| PSQI (SD) <sup>a</sup> | 5.37 (3.30) | 5.57 (3.30) | 5.28 (3.16) | 5.07 (3.70) | 4.22 (3.64) | 4.25 (2.73) | 0.0025 | MCI+V, AD, AD+V < CU, SCI, MCI |
| Self-reported sleep duration (SD) <sup>a</sup> | 6.94 (1.26) | 6.95 (1.17) | 7.15 (1.30) | 7.26 (1.47) | 8.07 (1.53) | 8.27 (2.03) | >0.00001 | CU, SCI, MCI, MCI+V < AD, AD+V |
| WMH burden, cm<sup>3</sup> (SD) <sup>a</sup> | 8.39 (6.12) | 10.8 (9.28) | 10.6 (8.27) | 29.8 (18.1) | 14.0 (10.3) | 32.5 (17.9) | >0.00001 | CU < SCI, MCI < AD < MCI+V, AD+V |
| Presence of infarcts, % (N) <sup>b</sup> | 11% (12) | 16% (21) | 9% (22) | 52% (68) | 15% (13) | 42% (18) | >0.00001 | CU, SCI, MCI, AD < MCI+V, AD+V |
| Presence of microbleeds, % (N) <sup>b</sup> | 15% (16) | 19% (25) | 18% (42) | 29% (39) | 17% (15) | 35% (15) | 0.0070 | n.s. |
| Probable or possible CAA, % (N) <sup>b</sup> | 12% (13) | 16% (22) | 17% (39) | 26% (34) | 16% (14) | 30% (13) | 0.0259 | n.s. |
| Geriatric Depression Scale (SD) <sup>a</sup> | 3.20 (3.89) | 4.84 (3.80) | 6.85 (5.19) | 6.43 (4.79) | 5.89 (4.59) | 5.56 (4.47) | >0.00001 | CU < SCI, MCI, MCI+V, AD, AD+V;<br>SCI < MCI, MCI+V |
| Geriatric Anxiety Inventory (SD) <sup>a</sup> | 2.85 (3.98) | 2.45 (2.65) | 4.00 (4.19) | 3.40 (3.75) | 2.80 (3.59) | 2.95 (3.99) | 0.0057 | CU, SCI, AD < MCI |
| MoCA (SD) <sup>a</sup> | 27.7 (1.65) | 27.3 (1.90) | 23.5 (3.01) | 23.1 (3.40) | 18.5 (4.26) | 18.0 (3.46) | >0.00001 | AD, AD+V < MCI, MCI+V < CU, SCI |

### 3.2 Non-linear relationship between sleep duration and WMH burden

When modelling the non-linear relationship between sleep duration and WMH burden within each group, neither the linear nor non-linear model reached statistical significance in explaining WMH burden (Figure 2A). However, when analyzing all participants together, the linear model significantly explained variance in WMH burden (Figure 2B: P linear = 0.003), and the non-linear model provided a significantly better fit than the linear one (Figure 2B: P non-linear = 0.002; Supplementary Table 1).

**Figure 2.**
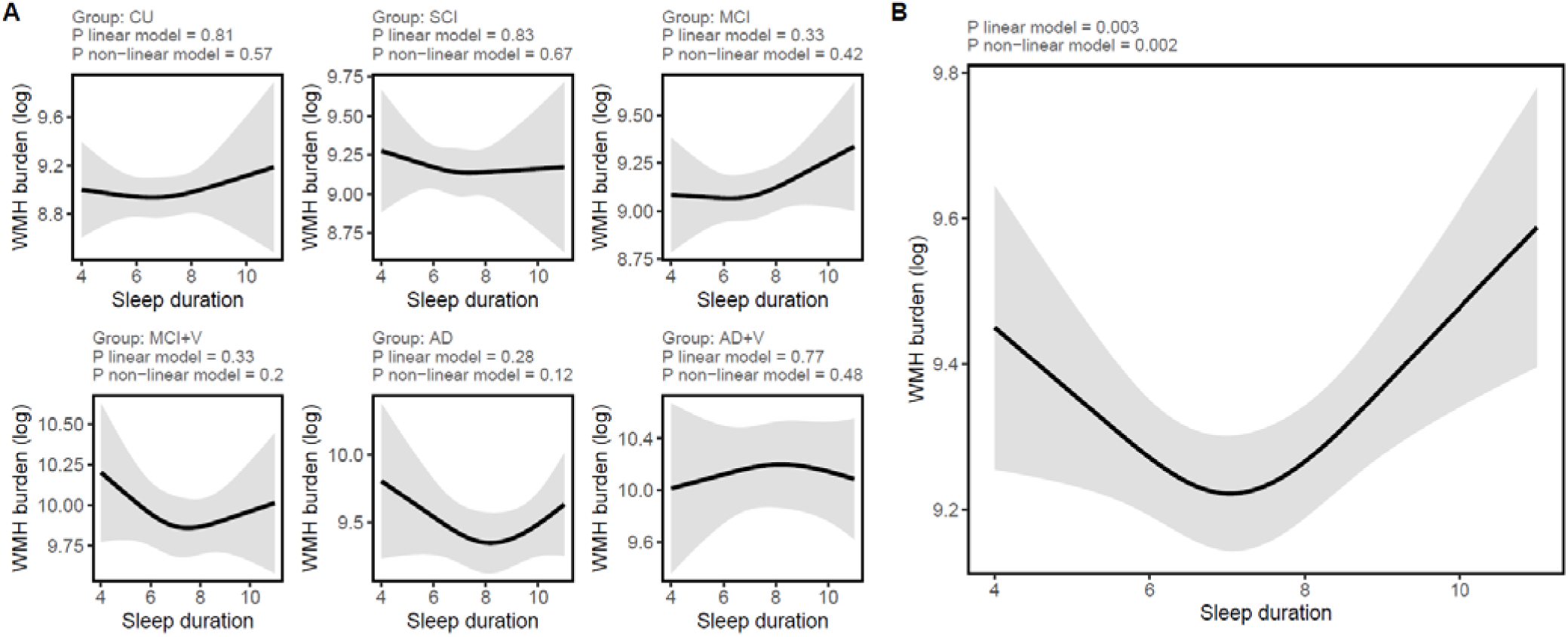
Non-linear relationship between WMH burden and sleep duration for (A) each diagnostic group, and (B) across all participants.

### 3.3 Relationship between sleep duration and WMH+ status

We did not observe significant differences in self-reported sleep duration between WMH+ and WMH-only subgroups (Supplemental Figure 2).

### 3.4 Relationship between WMH burden, sleep duration, and WMH+ status

A significant three-way interaction was observed between sleep duration, WMH+ status, and diagnostic group (Table 2, Figure 3), with significant effects in MCI+V [b = −0.26, p = 0.05, SE = 0.13] and AD [b = −0.35, p = 0.018, SE = 0.15]. Among WMH+ individuals, shorter sleep duration was associated with greater WMH burden in MCI+V and AD, and this slope was significantly amplified relative to CU. No significant associations were observed in the CU reference group. A significant two-way interaction between WMH+ status and diagnostic group was also detected, with a significant contrast in AD [b = 2.54, p = 0.023, SE = 1.12], indicating greater WMH burden in WMH+ relative to WMH-only in this group. Age showed a significant main effect [b = 0.04, p < 0.0001, SE = 0.003]. Adjustment for sleep breathing disorder did not alter the observed three-way and two-way interactions (Supplementary Table 2).

**Figure 3.**
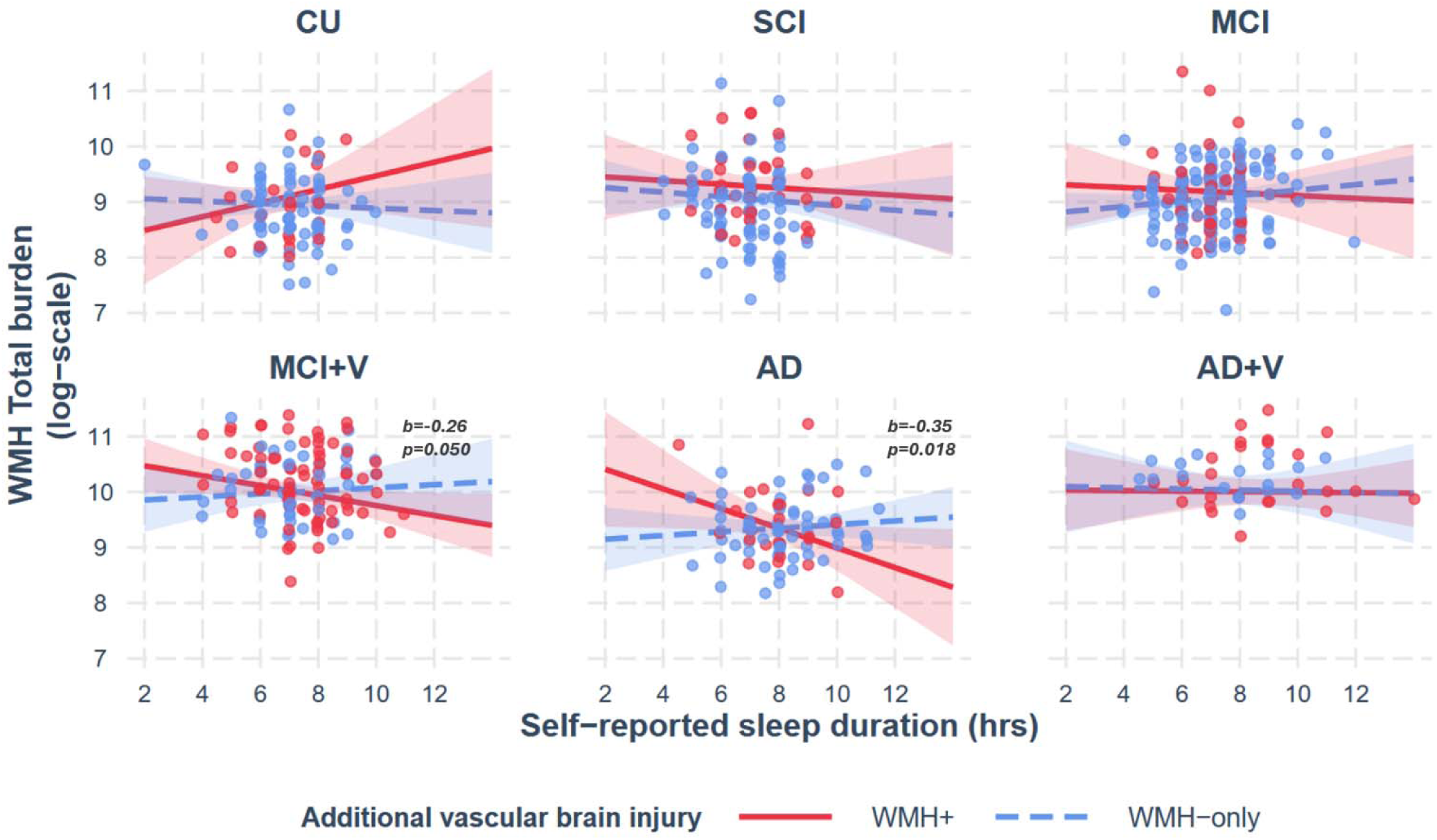
The relationship between WMH burden and self-reported sleep duration is amplified with WMH+ status in the MCI+V and AD groups. The significant associations reflect the differential slope in MCI+V or AD individuals compared to the CU reference group, where no associations were found.

**Table 2.**
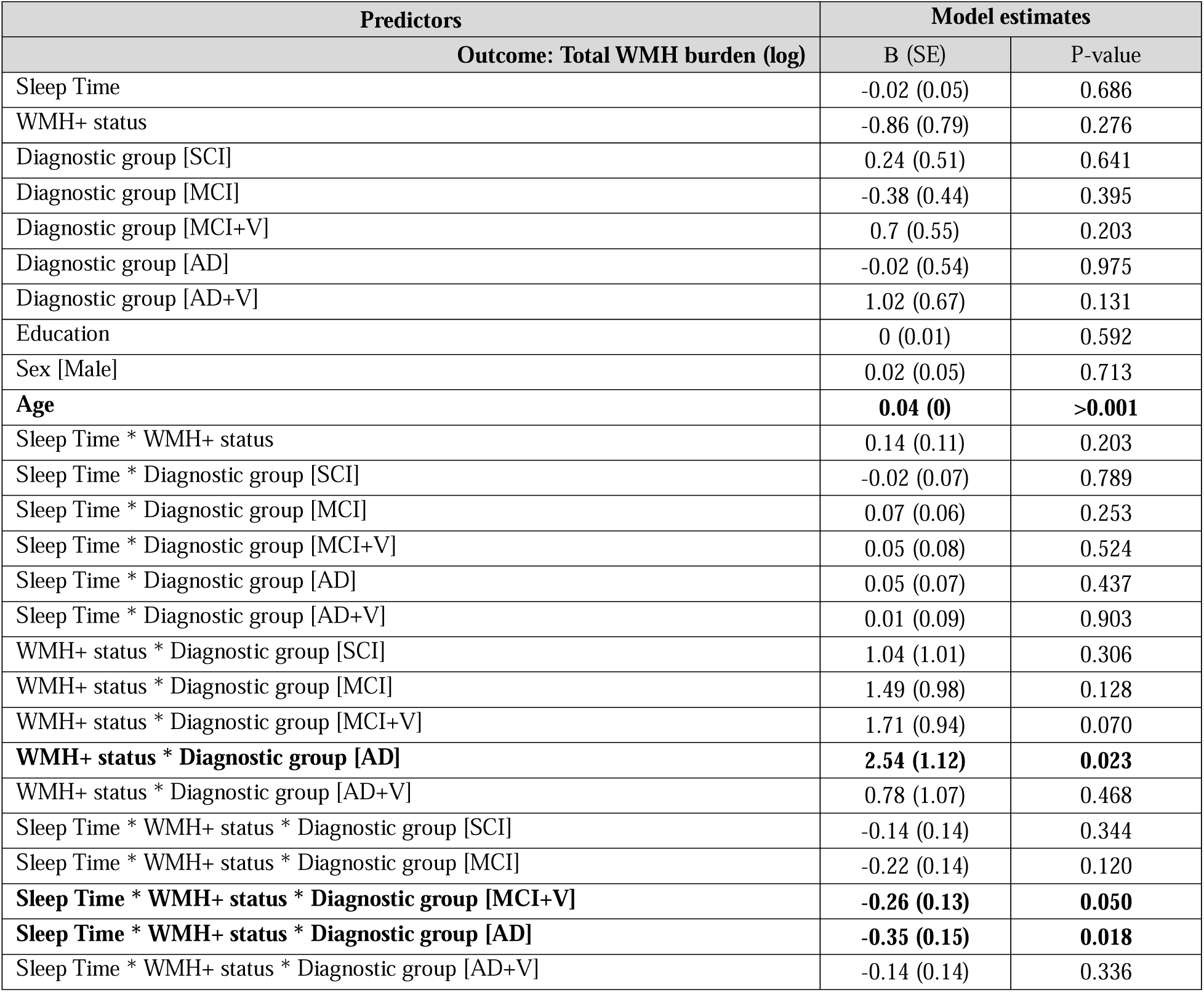
Model 1: Total WMH burden (log) ∼ Sleep Time * WMH+ status * Group + Age + Sex + Education. WMH+ status is dummy-coded (0 = WMH-only), such that the coefficients represent the differences relative to individuals without additional burden (WMH-only vs WMH+). Group is coded as a factor, with CU set as the reference. Sex is coded as a factor, with Female set as the reference.

When modeled using the more complex four-level vascular brain injury variable, the three-way interaction remained significant in MCI+V and AD, and was additionally observed in MCI [b = −0.143, p = 0.039, SE = 0.07; Supplementary Table 3].

#### 3.4.1 Relationship between WMH burden, PSQI scores and WMH+ status

We did not find a significant relationship between either total score on the PSQI or scores for each sub-domain of the PSQI (excluding sleep duration) with WMH burden, nor did we find an interaction with additional vascular brain injury (Table 3). However, we found a significant association between the daytime disturbances sub-domain and diagnostic group for the SCI group (b=0.48, p=0.01; Supplementary Table 4), such that there is a stronger positive association between WMH burden and daytime disturbances in the SCI group when compared to the CU reference group.

**Table 3.** Results from linear regressions between PSQI sub-domain scores and WMH burden. Age, Sex and Education are included as covariates in all models. WMH+ status is dummy-coded (0 = WMH-only, 1 = WMH+), such that the coefficients represent the differences relative to individuals without additional burden. Adj. p-value: Adjusted p-value following FDR correction for six models. The full model output can be found in the Supplementary Material.

| Outcome: Total WMH burden (log) | Model estimates |  |
| --- | --- | --- |
|  | B (SE) | Adj. P-value |
| Model 1: |  |  |
| PSQI total score | -0.01 | 0.87 |
| PSQI total score * WMH+ status | -0.01 | 0.93 |
| Model 2: |  |  |
| PSQI Sleep quality score | -0.06 | 0.79 |
| PSQI Sleep quality score * WMH+ status | 0.08 | 0.80 |
| Model 3: |  |  |
| PSQI Sleep latency score | -0.03 | 0.92 |
| PSQI Sleep latency score * WMH+ status | -0.01 | 0.99 |
| Model 4: |  |  |
| PSQI Sleep efficiency score | -0.07 | 0.71 |
| PSQI sleep efficiency score * WMH+ status | -0.10 | 0.37 |
| Model 5: |  |  |
| PSQI Sleep medication score | 0.02 | 0.90 |
| PSQI Sleep medication score * WMH+ status | 0.15 | 0.12 |
| Model 6: |  |  |
| PSQI Sleep disturbances score | 0.24 | 0.42 |
| PSQI Sleep disturbances * WMH+ status | -0.18 | 0.56 |
| Model 7: |  |  |
| PSQI Daytime disturbances | -0.21 | 0.32 |
| PSQI Daytime disturbances * WMH+ status | 0.04 | 0.91 |

### 3.5 Relationship between cognitive performance, sleep duration and WMH+ status

A significant three-way interaction between WMH+ status, sleep duration, and diagnostic group was observed for measures of attentional processing and executive function in the AD group (Figure 4; TMT-B: b= 2.19, p=0.01, p-adj=0.12; Stroop: b= 2.32, p=0.01, p-adj=0.11; CRT: b=0.59, p=0.01, p-adj=0.11; Supplementary Tables 5–10). Similar three-way interactions were detected in the AD+V group for memory and executive measures (RAVLT: b=0.71, p=0.03, p-adj=0.21; Stroop: b=2.47, p=0.03, p-adj=0.21). However, these effects did not remain significant after FDR correction.

**Figure 4.**
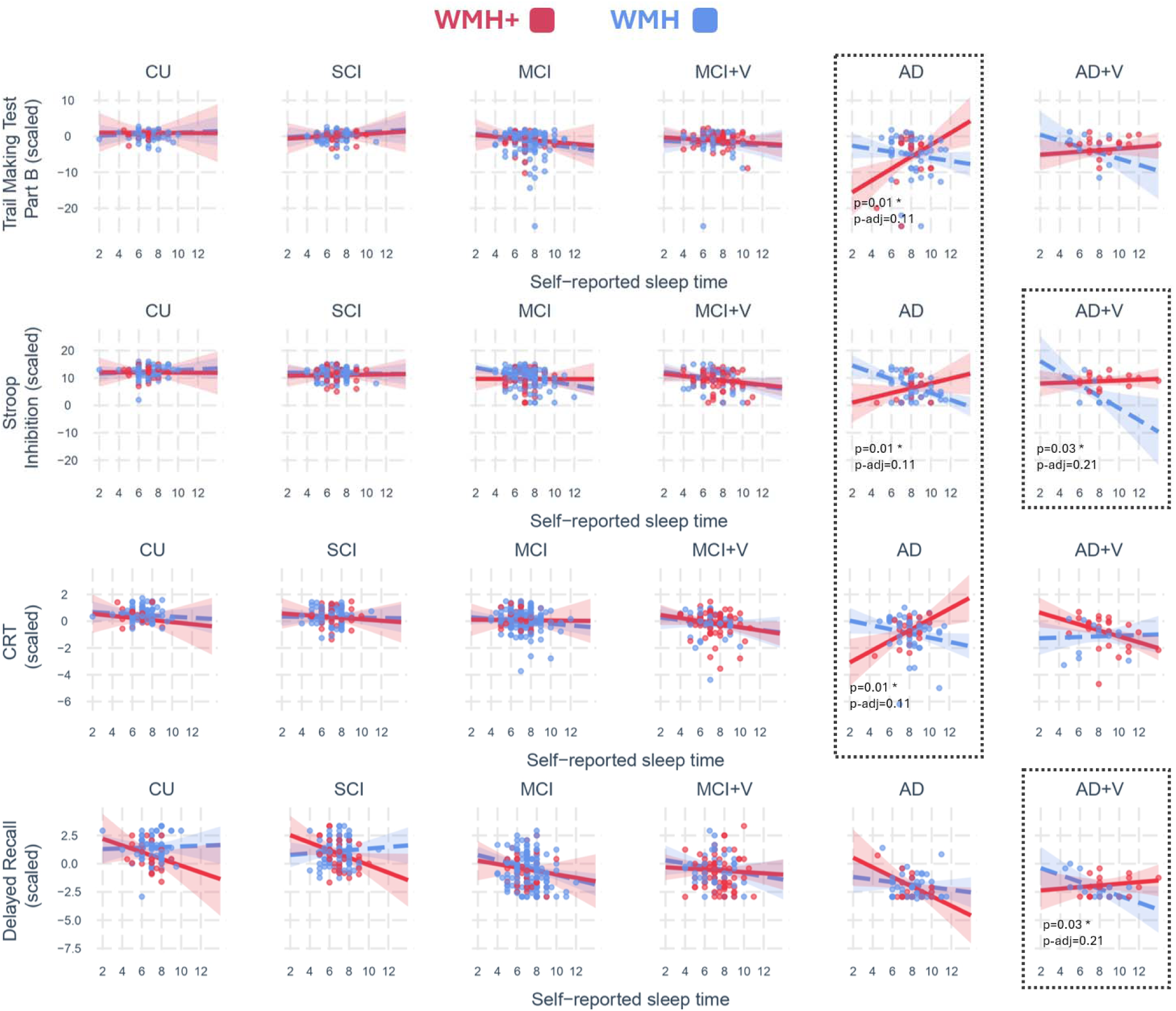
Significant relationships between cognitive performance and self-reported sleep time, stratified by WMH+ status, across diagnostic groups. Shaded regions reflect 95% confidence intervals. P-values reflect three-way interactions (sleep duration * vascular brain injury burden * diagnostic group) from linear models; adjusted p-values are FDR-corrected for 6 comparisons. Performance was reverse scored when needed, such that higher values reflect better performance.

Exploratory analyses, using post-hoc simple slope analysis, revealed a consistent directional pattern across cognitive tests. Among AD individuals with WMH+, shorter sleep duration was associated with worse executive and processing speed performance (TMT-B: b=1.66, p=0.003; CRT: b=0.40, p=0.006). In contrast, among those with WMH-only, longer sleep duration was associated with poorer executive performance. A comparable pattern was observed in AD+V, where, in WMH-only individuals, longer sleep was associated with worse performance on the Stroop [b=-2.14, p=0.015] and showed a trend toward worse delayed recall performance [b=-0.30, p=0.055]. No significant associations between sleep and cognition were observed in the CU reference group.

We next examined whether these two AD subgroups (i.e., WMH-only and WMH+) differed in demographic characteristics, overall cognitive function, neuropsychiatric characteristics or fluid biomarkers of AD pathology (Supplementary Table 11). We found that both subgroups were comparable across most variables, with the exception that individuals with WMH+ were older (p=0.03) and showed greater decline on the MoCA after 2-year follow-up (p=0.05).

#### 3.5.1 Prediction of future cognitive function using sleep duration and WMH+ status at baseline

Of the 105 CU and 133 SCI participants, only 12 CU individuals changed diagnosis at follow-up (MCI: n=8; MCI+V: n=1; SCI: n=3). Among the SCIs, 15 progressed to MCI (MCI: n=12; MCI+V: n=3) and 28 reverted to CU. Given the small number of participants who progressed to worse cognitive status, we predicted change in cognitive performance across all participants rather than modelling conversion within the CU groups only.

When predicting change in MoCA scores (follow-up performance minus baseline), we did not find a significant association between baseline self-reported sleep duration, nor any interactions with groups or WMH+ (Supplementary Tables 12-17). Amongst the other cognitive measures, a significant two-way interaction was observed between baseline self-reported sleep duration and diagnostic group when predicting change in Stroop performance for AD [b=1.19, p=0.017, p-adj=0.99]. More specifically, relative to the CU group, longer baseline self-reported sleep duration in AD was associated with a greater decline in Stroop performance at follow-up. However, this did not survive FDR corrections.

## 4. Discussion

Sleep is increasingly recognized as a modifiable factor linked to cognitive decline^31,32^, though it may also signal underlying disease processes. How it interacts with vascular brain injury – an important contributor to AD^2,12,52^ – remains unclear. We hypothesized that self-reported sleep duration would be longer in WMH+ individuals and that, if longer sleep indexes more severe pathology, WMH+ would amplify the relationship between longer sleep and WMH burden. Contrary to expectations, WMH+ did not relate to longer sleep duration per se, but instead modified the relationships among sleep duration, WMH burden, and cognition across the AD continuum. Specifically, in MCI+V and AD groups, WMH+ amplified the relationship between shorter sleep and greater WMH burden – a pattern that was also observed in our more complex model of additional vascular brain injury, where effects also emerged in MCI. In AD dementia (AD, AD+V), vascular brain injury profile further differentiated patterns: among WMH+ individuals, shorter sleep was associated with greater WMH burden and worse cognition, whereas in those with WMH-only, longer sleep was associated with worse cognition. No significant associations were observed in CU and SCI individuals, suggesting the interactions between sleep and vascular brain injury may emerge primarily in symptomatic stages.

These insights were made possible by a unique and deeply phenotyped pan-Canadian cohort spanning CU to AD dementia. By focusing on clinically defined vascular groups in symptomatic participants (MCI+V, AD+V), and applying a composite measure of microbleeds, infarcts, and probable or possible CAA across the full cohort, we captured cerebrovascular pathology along a continuum from primary to secondary contributors to cognitive decline. Notably, WMH+ modified cross-sectional associations, but baseline vascular brain injury did not interact with sleep to predict cognitive change over the 2-year follow-up, a timeframe that may limit detection of longer-term effects. By demonstrating that vascular brain injury can shape the impact of sleep in AD, these findings highlight the importance of accounting for cerebrovascular comorbidity when interpreting sleep’s effect on cognition, ultimately informing its use as a potential biomarker or therapeutic target.

Our findings expand on the limited research examining sleep and vascular brain injury in the AD continuum. While sleep disturbances are well documented in AD^53–55^, very few have explicitly investigated the intersection of sleep, vascular brain injury, and AD^26,56^. One study included participants with AD and MCI, but was limited to broadly defined sleep disturbances and WMHs^56^. Another study included amyloid PET, alongside self-reported sleep duration and WMHs, but only in individuals free of dementia^26^. Beyond these two studies, research on sleep and vascular brain injury has largely focused on CU adults, often using a single neuroimaging marker of vascular brain injury (i.e., WMHs)^23,30,56^. By directly modeling cerebrovascular pathology in addition to WMHs across the AD continuum, our study provides novel insight into how WMH+ modifies the relationships between sleep, WMH burden, and cognitive outcomes.

### 4.1 Non-linear relationship between sleep duration and WMH burden

While we did not observe a non-linear relationship between sleep duration and WMH burden when investigating each group, we observed one when modelling all participants together. This U-shape replicates previous findings and places the optimal sleep duration ^22,23^, with the lowest amount of WMH burden, around 7 hours. The fact that the U-shape appeared only in the combined cohort may indicate that different diagnostic groups occupy distinct positions along this curve, reflecting group-specific vulnerabilities or compensatory mechanisms. These findings raise the possibility that deviations from optimal sleep duration could interact with underlying vascular and neurodegenerative processes differently across the cognitive spectrum. Future work using longitudinal and objective sleep measures will be essential to determine whether modifying sleep duration could influence WMH accumulation or slow cognitive decline.

### 4.2 Amplified relationship between shorter sleep and higher WMH burden in individuals with WMH+

We had hypothesized that WMH+ would amplify the relationship between longer sleep and greater WMH burden. Contrary to this, we observed an amplified relationship between shorter sleep and greater WMH burden in WMH+ individuals. Because sleep duration is continuous, it is difficult to determine whether this reflects a harmful effect of shorter sleep or a potential benefit of longer sleep. Experimental evidence supports the former: chronic sleep fragmentation induced white matter abnormalities in an animal model of cerebral small vessel disease, but not in controls, suggesting that cerebrovascular pathology increases susceptibility to disturbed sleep^57^. The fact that this pattern was observed in the AD and MCI+V groups — but not in AD+V — may reflect the small sample size in the AD+V group, and replication is needed.

### 4.3 WMH+ modifies the direction of the relationship between sleep and cognition in AD dementia

Exploratory analyses on cognition revealed a consistent directional divergence in AD dementia (AD, AD+V) by WMH+ status, although these interactions did not survive correction for multiple comparisons. Among participants with WMH+, shorter sleep was associated with worse cognition, while longer sleep was associated with worse cognition in WMH-only participants. This contrasts with our original hypothesis that WMH+ would amplify the relationship between longer sleep and cognitive impairment. Interestingly, we observe patterns that mirror the two prevailing hypotheses in the literature: longer sleep as a potential compensatory mechanism, and as a marker of underlying neurodegenerative burden. These two patterns may reflect distinct underlying mechanisms that may depend on the overall burden of vascular brain injury.

In WMH+ individuals, the association between shorter sleep and worse cognition aligns with prior work proposing a compensatory role for longer sleep in at-risk populations, even in the presence of elevated inflammatory markers^20^. In WMH-only individuals, shorter sleep was associated with better cognition – aligning with the hypothesis that longer sleep reflects underlying neurodegeneration ^58^ as opposed to compensation. Some studies suggest AD may be characterized by a hypersomnia phenotype, with both frequent daytime napping^59^ and excessive daytime sleepiness^60^ being associated with preclinical AD, and post-mortem work showing tau-related degeneration of wake-promoting neurons^61^.

It is possible that both mechanisms, either compensatory or reflecting underlying neurodegeneration, exist across different individuals within the AD continuum and may reflect different stages of disease progression, or different AD subgroups. However, when we compared our own AD subgroups (i.e., WMH vs WMH+) across demographic, clinical and fluid biomarker characteristics, we did not observe any significant differences other than the WMH+ group being significantly older. This suggests that additional vascular brain injury burden itself may be the key variable that differentiates across these subgroups, and might be the main driver shaping the association between sleep and cognition. Importantly, similar self-reported sleep duration may mask differences in the underlying sleep architecture. It is well established that the architecture of sleep is altered in AD^54^, characterized by an increase in lighter non-REM sleep, a decrease in slow-wave sleep and REM sleep, and greater sleep fragmentation. As such, our two AD subgroups (WMH-only vs WMH+) may differ in the underlying architecture of their sleep, beyond what self-reported duration captures. Given evidence that sleep architecture varies across AD phenotypes^62^, such differences may contribute to the heterogeneous associations we observe with cognition.

### 4.4 Limitations

While our study has several strengths, several limitations merit consideration. Our sample size, while reasonable, was insufficient to stratify WMH-only and WMH+ subgroups by short and long sleepers. Quantitative measures were available for WMHs, but not for infarcts or microbleeds, limiting exploration of more nuanced vascular contributions. The multiple interaction terms included may have reduced statistical power, potentially contributing to null findings. Longitudinal analyses were constrained by a relatively short follow-up (∼2 years) and low conversion rates from CU and SCI to MCI. Finally, our study relied on self-reported sleep, which can differ from objectively measured sleep, particularly in older adults and those with cognitive impairment^63,64^. While the PSQI is widely used in large cohort studies linking sleep to dementia risk and vascular brain injury^14–17,23^, it is a general screen for sleep disturbances rather than a tool to capture the precise sleep dimensions most relevant to cognition. Despite these caveats, our results consistently show that vascular brain injury shapes the relationship between sleep and cognition. These findings emphasize the importance of comprehensive vascular assessment in studies of sleep and cognitive aging, and point to future opportunities to refine these relationships using objective sleep measures, more detailed vascular metrics, and longer follow-up.

## Conclusion

Our results highlight the importance of considering vascular brain injury when examining the relationship between sleep and both WMH burden and cognition in AD. While sleep is a modifiable factor, the mechanisms linking it to dementia remain unclear – importantly these mechanisms are unlikely to be uniform, and may vary depending on factors, such as vascular brain injury. Since self-reported sleep duration is readily available in multiple large cohorts, our results can easily be replicated. Replication will also help confirm whether the heterogeneous associations observed with cognition in AD, depending on the severity of vascular brain injury, hold across different cohorts. As vascular contributions to AD are increasingly recognized, integrating them in the study of sleep in AD is essential, if we aim to effectively leverage sleep as either a biomarker or a target for intervention

## Data Availability

Data used in preparation of this article were obtained from the Comprehensive Assessment on
Neurodegeneration and Dementia (COMPASS-ND) study and are available on request from the corresponding author and upon approval by the Canadian Consortium on Neurodegeneration in Aging.

## ACKNOWLEDGEMENT

The authors would also like to thank all the participants in this study for their time.

## CONFLICT OF INTEREST STATEMENT

All authors declare that they have no financial, personal, or competing interests/conflicts.

## CONSENT STATEMENT

Ethical approval was obtained from the aging-neuroimaging research ethics committee of the CIUSSS du Centre-Sud-de-île-de-Montréal (CIUSSS-CSMTL), and from the McGill University Institutional Review Board, Faculty of Medicine and Health Sciences. Written informed consent was obtained from all participants.

## FUNDING SOURCES

This work was supported by the Fonds de Recherche Québec – Santé (FRQS), Fonds de soutien à la recherche pour les neurosciences du vieillissement from the Fondation Courtois, FRQS Chercheurs boursiers Junior 1 and 2, and the (A.B.); and postdoctoral scholarship from the Vascular Training Platform (S.L.). The Canadian Consortium on Neurodegeneration in Aging is supported by a grant from the Canadian Institutes of Health Research (CNA-137794 for CCNA Phase I; CNA-163902 for CCNA Phase II; CND-193575 for CCNA Phase III). ZI is funded by CIHR (BCA 527734), Gordie Howe CARES, and UK National Institute for Health and Care Research Exeter Biomedical Research Centre.

## Notes

### Competing Interest Statement

The authors have declared no competing interest.

### Funding Statement

This work was supported by the Fonds de Recherche Quebec - Sante (FRQS), Fonds de soutien a la recherche pour les neurosciences du vieillissement from the Fondation Courtois, FRQS Chercheurs boursiers Junior 1 and 2, and the (A.B.); and postdoctoral scholarship from the Vascular Training Platform (S.L.). The Canadian Consortium on Neurodegeneration in Aging is supported by a grant from the Canadian Institutes of Health Research (CNA-137794 for CCNA Phase I; CNA-163902 for CCNA Phase II; CND-193575 for CCNA Phase III). ZI is funded by CIHR (BCA 527734), Gordie Howe CARES, and UK National Institute for Health and Care Research Exeter Biomedical Research Centre.

### Author Declarations

The aging-neuroimaging research ethics committee of the CIUSSS du Centre-Sud-de-ile-de-Montreal (CIUSSS-CSMTL) gave ethical approval for this work. The McGill University Institutional Review Board of the Faculty of Medicine and Health Sciences gave ethical approval for this work.

