## Supplementary Files for "Vascular brain injury modifies the relationship between sleep duration, cognition, and white matter hyperintensity burden in the Alzheimer’s disease continuum"

### Supplementary Material

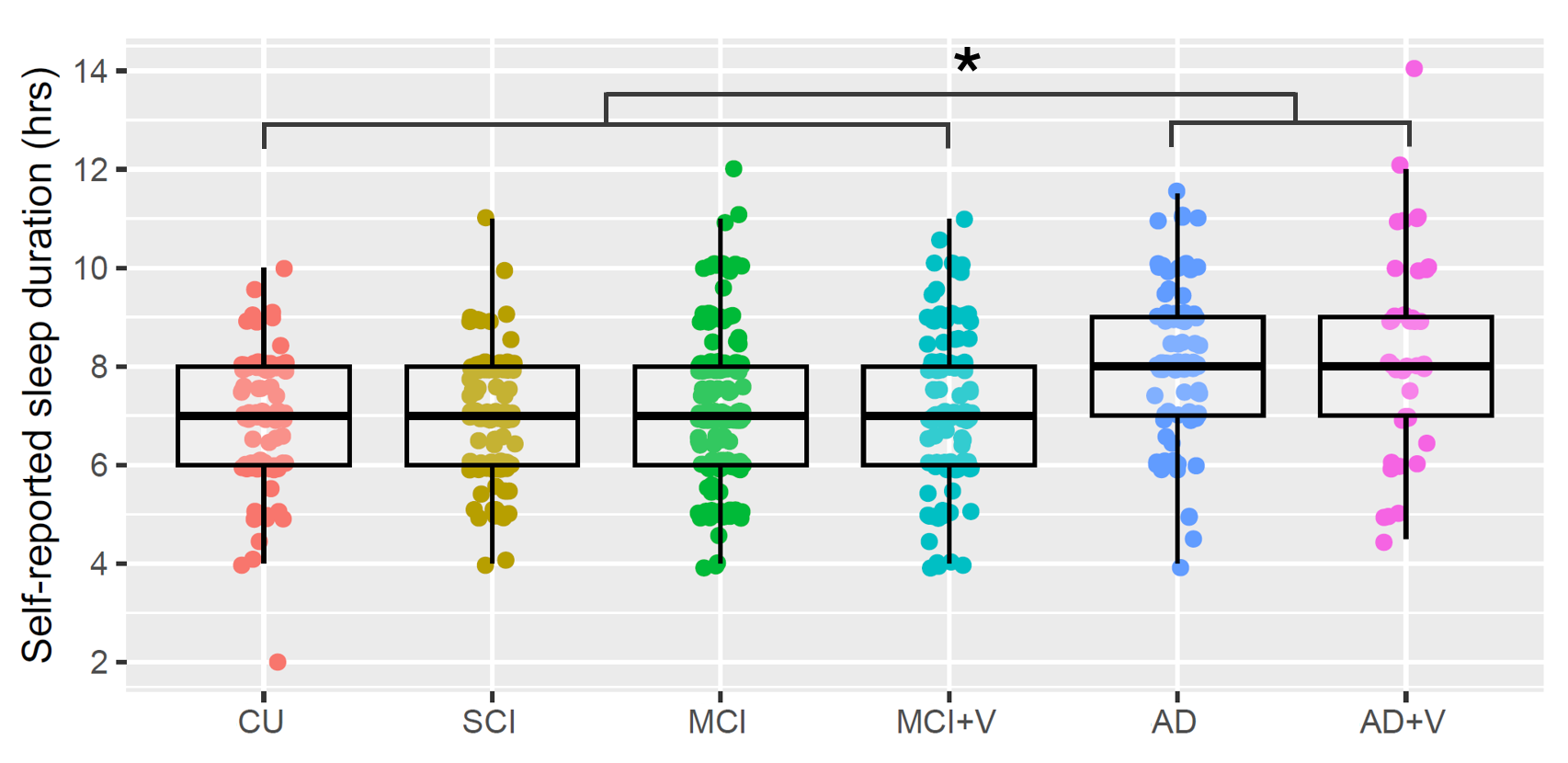

**Supplementary Figure 1. Self-reported sleep duration across diagnostic groups.** Asterisk denotes significant post-hoc pairwise Wilcoxon rank test comparison in self-reported sleep duration between AD, AD+V groups compared to CU, SCI, MCI and MCI+V (see Table 1).

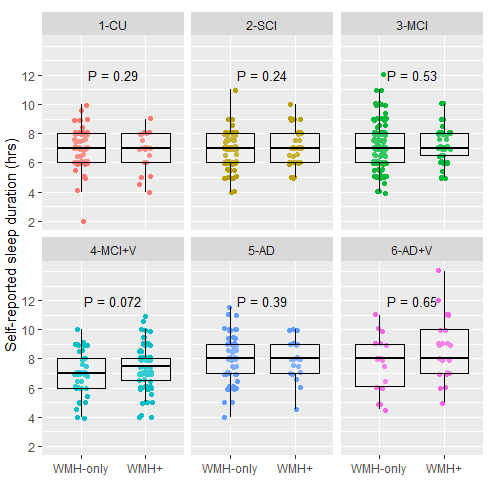

**Supplementary Figure 2.** Differences in self-reported sleep duration across the presence or absence of additional vascular brain injury burden, for each diagnostic group. P-value reflects statistical significance for group comparisons adjusted for Age, Sex and Education.

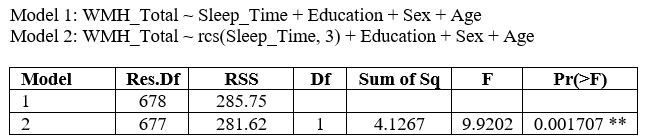

**Supplementary Table 1. Analysis of Variance comparing linear and non-linear modelling of Sleep Time.**

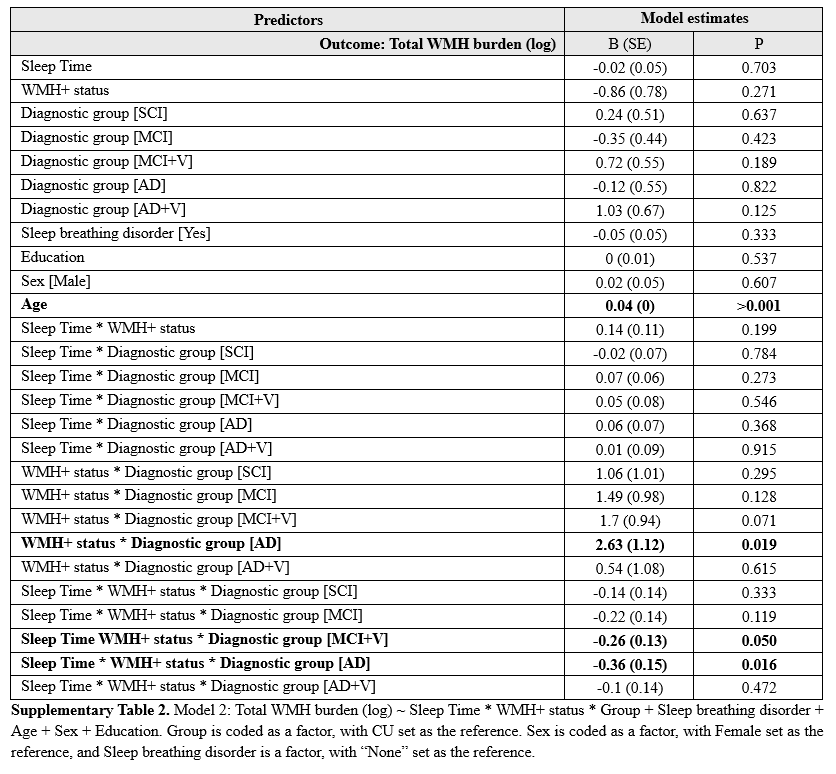

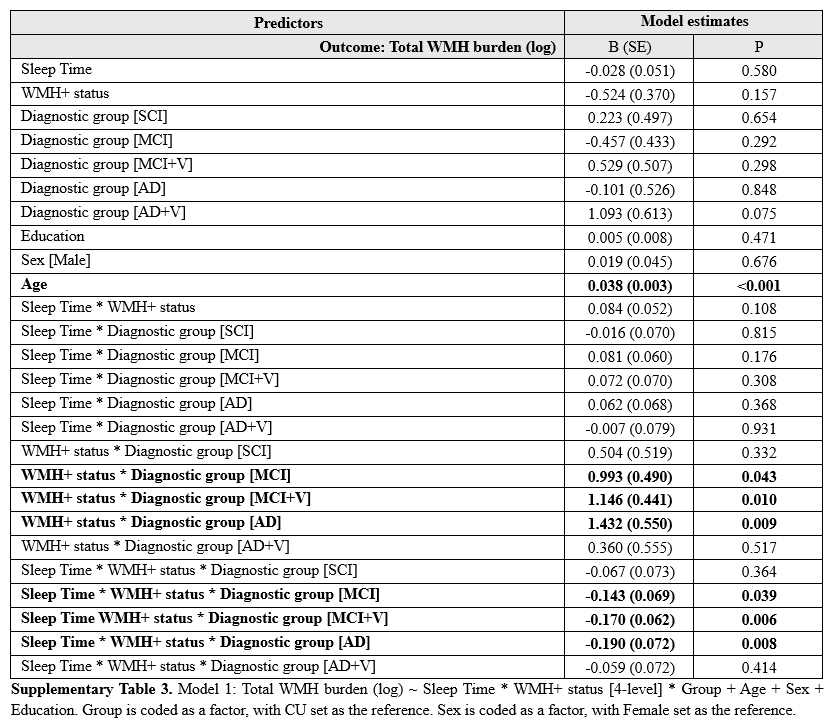

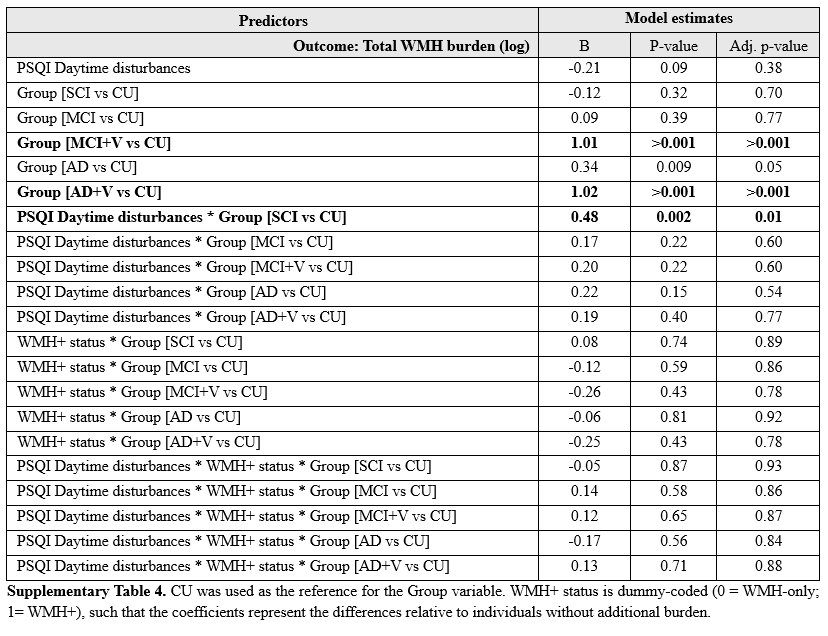

**Baseline cognition (FDR corrected):**

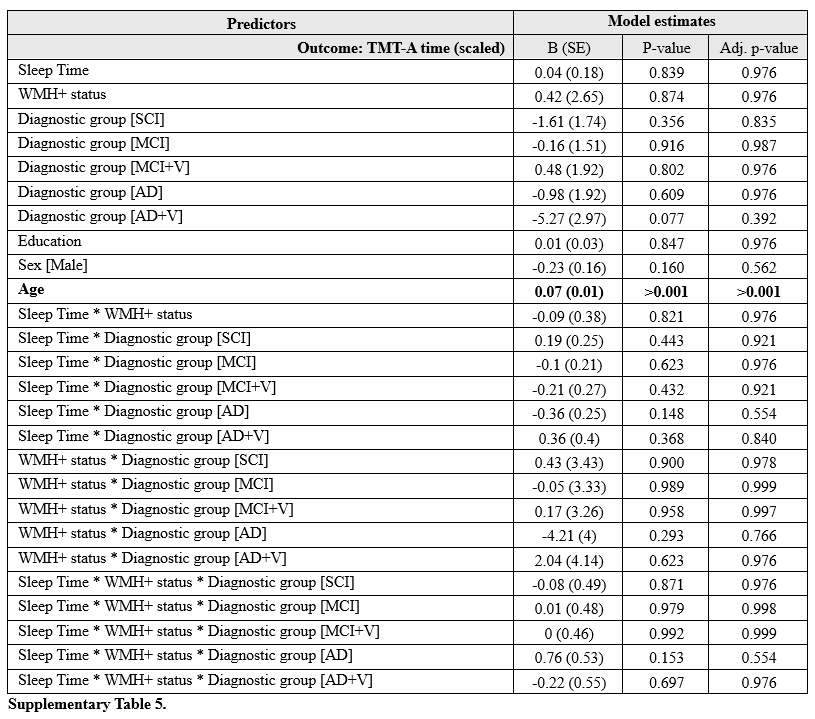

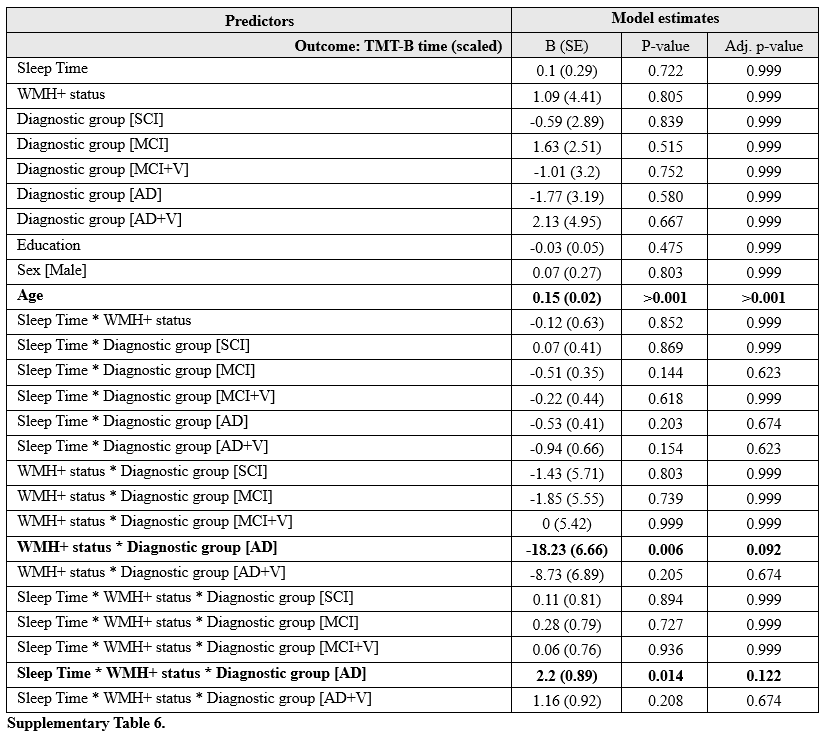

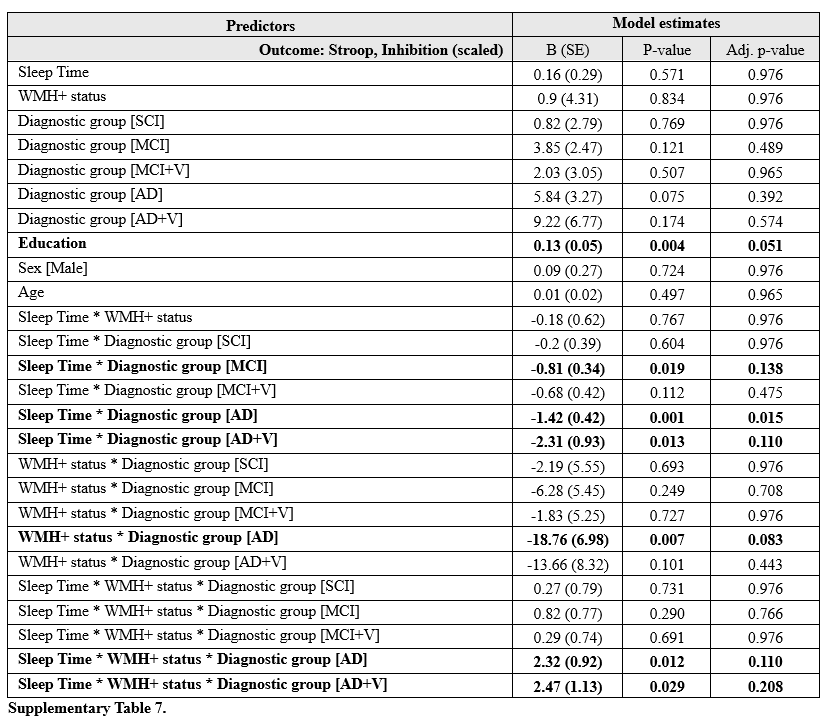

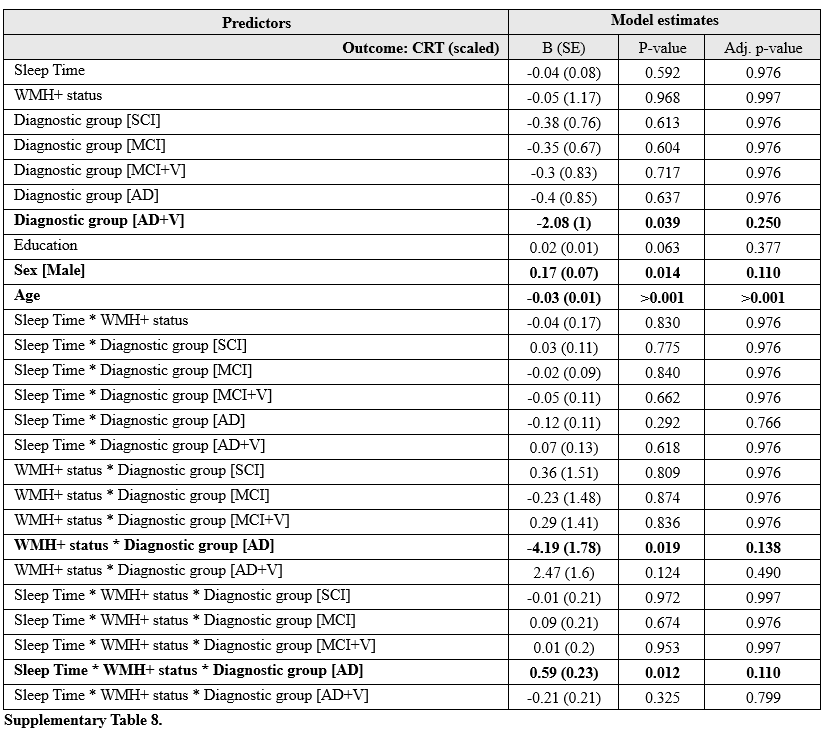

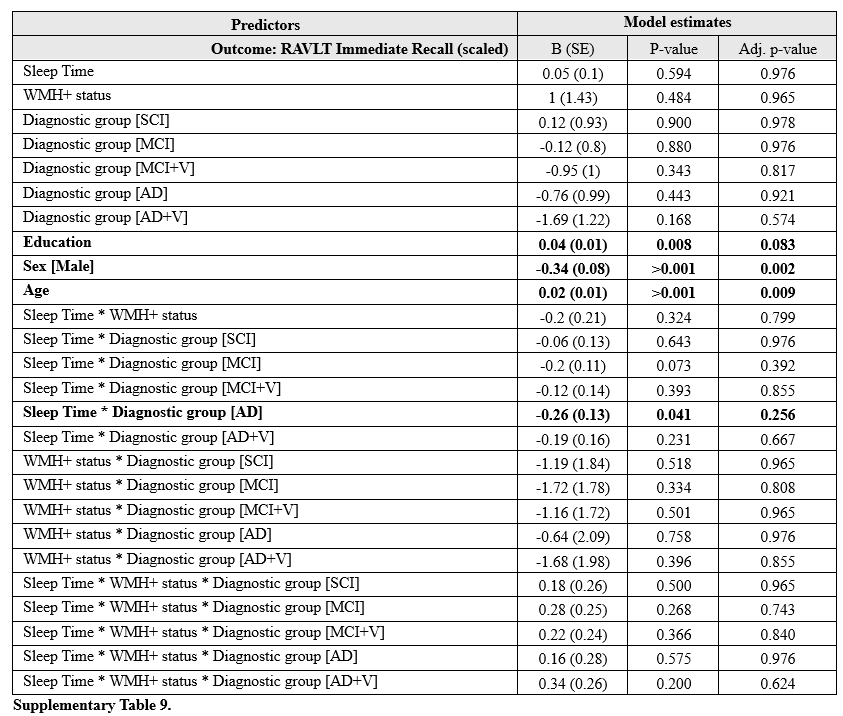

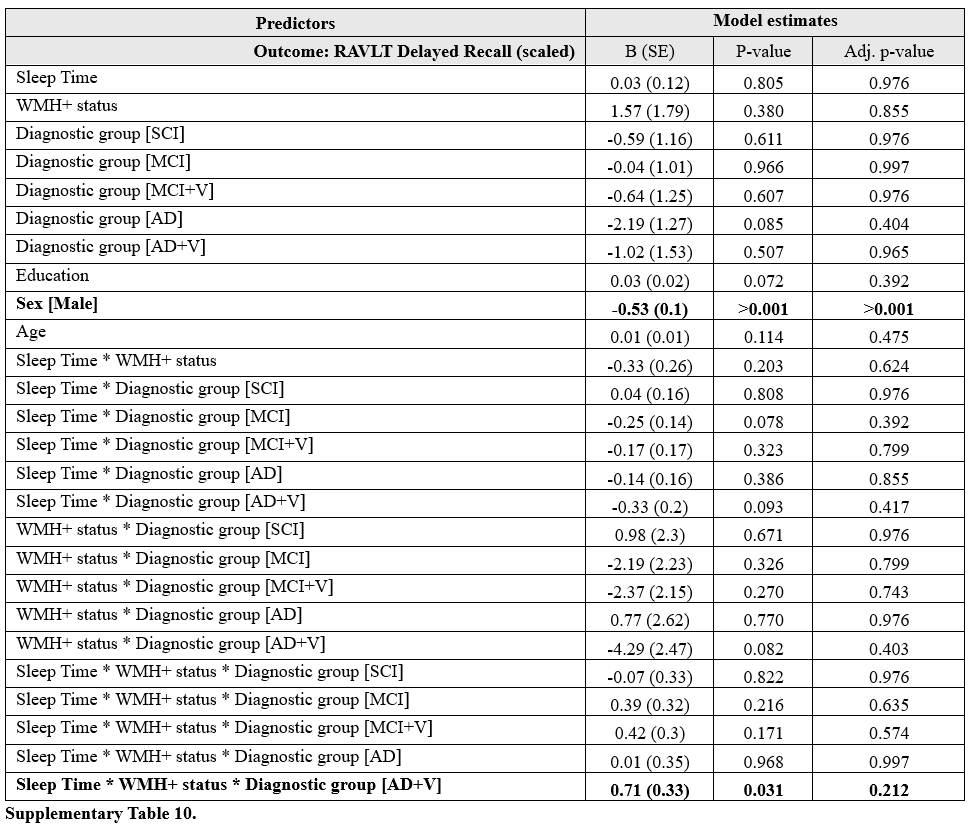

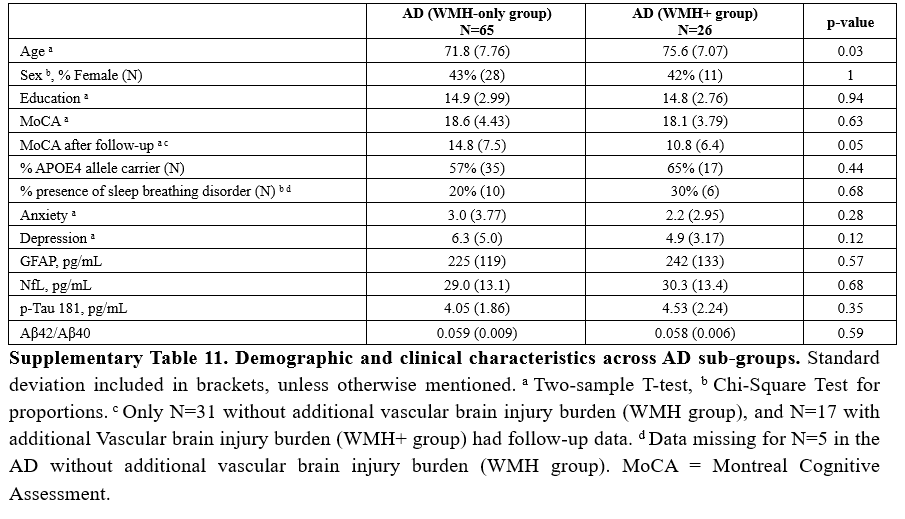

**Follow-up cognition Tables (FDR corrected):**

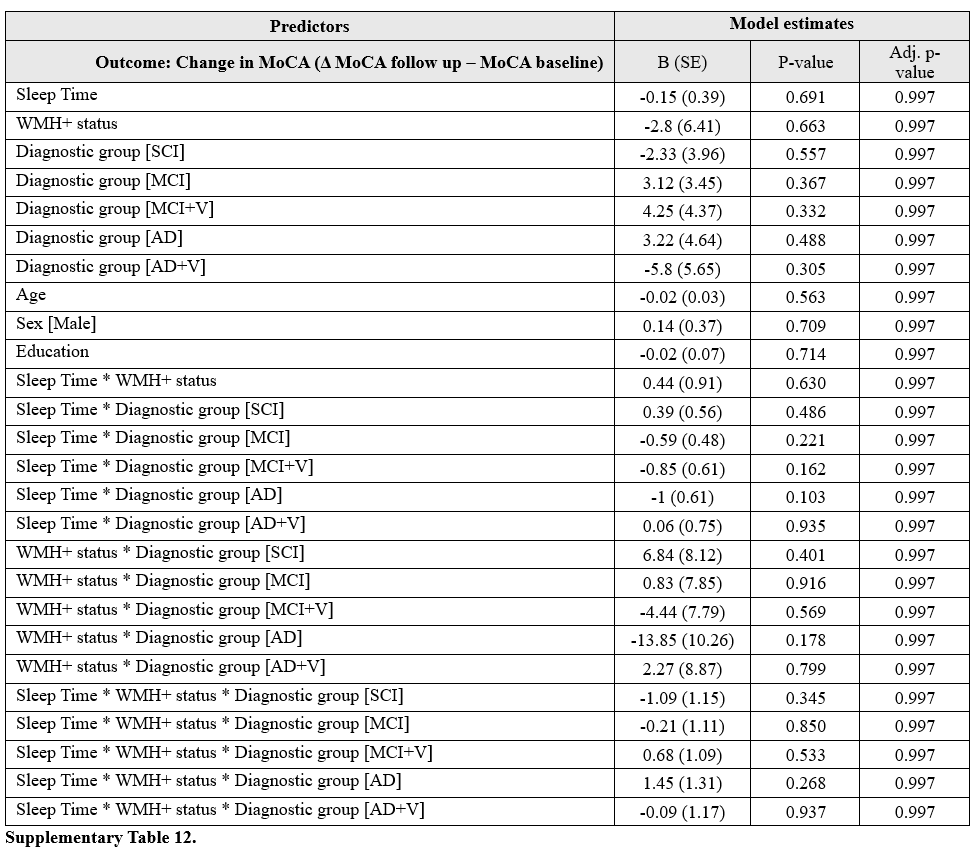

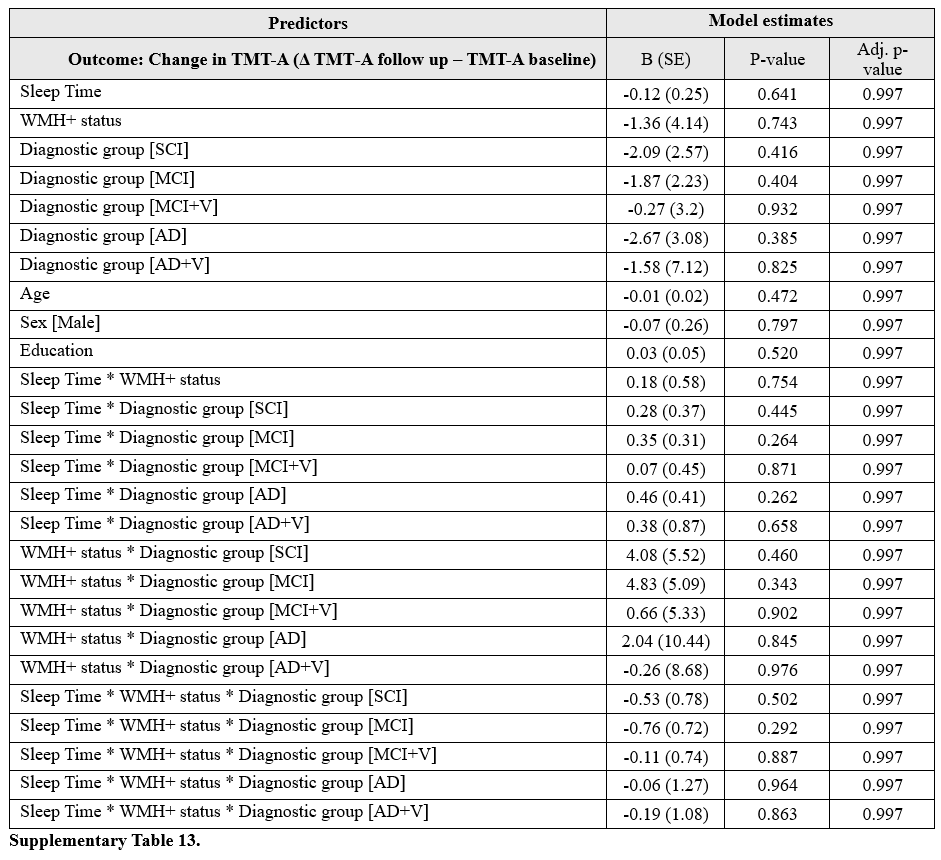

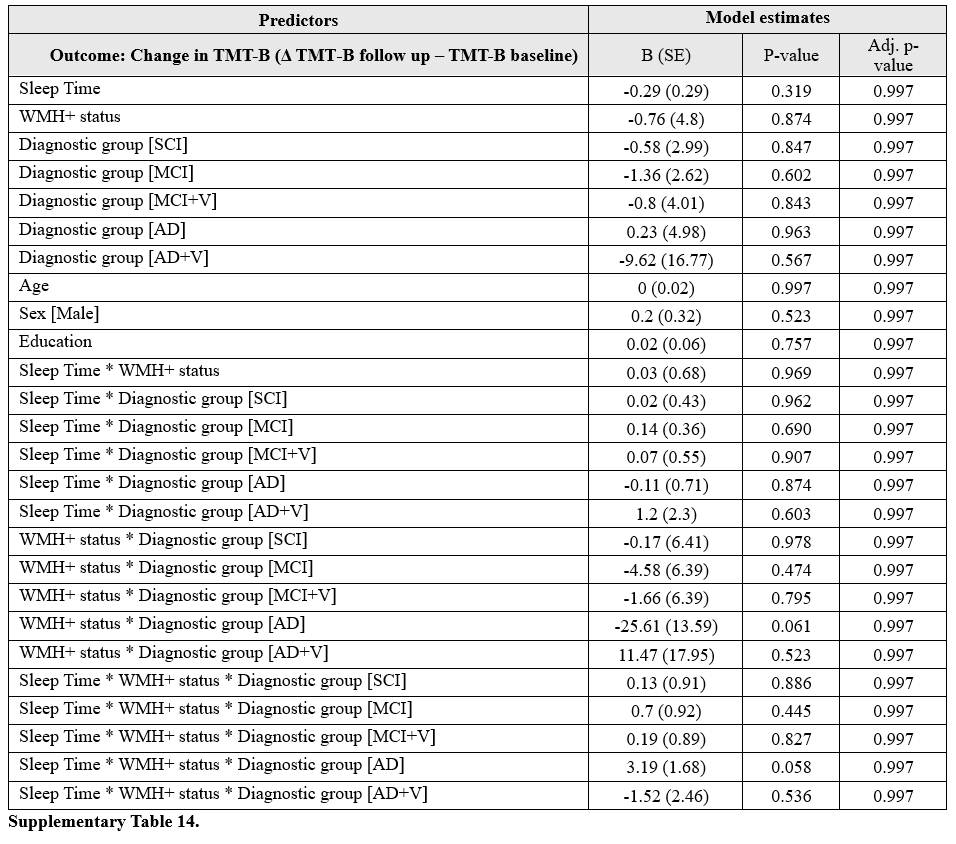

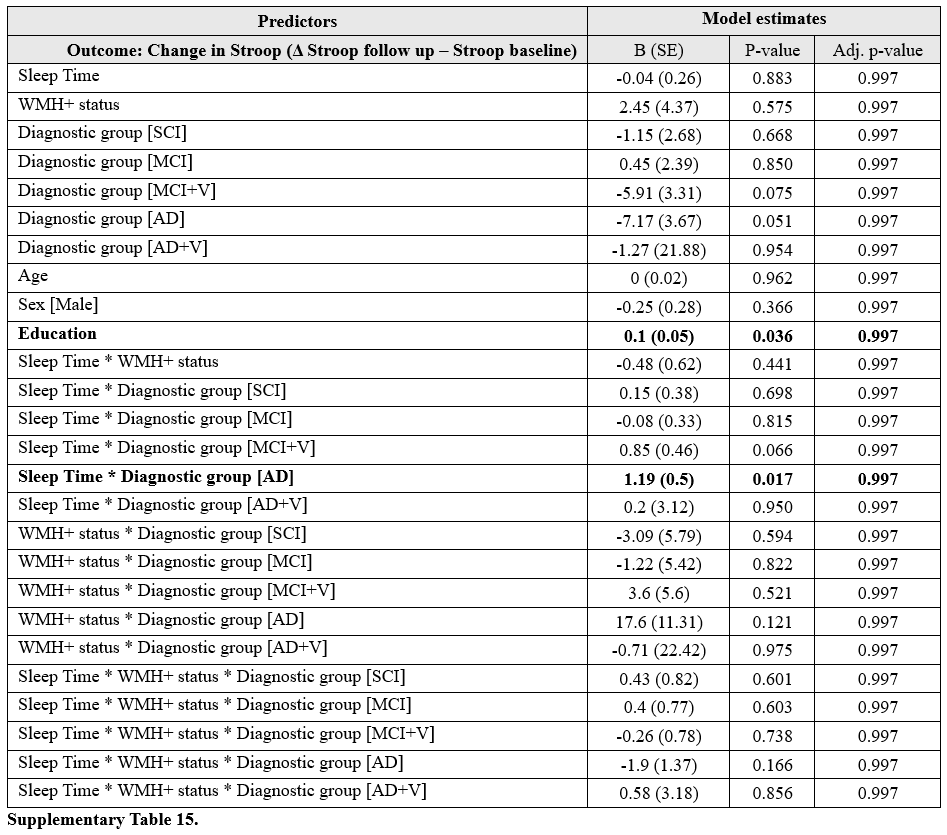

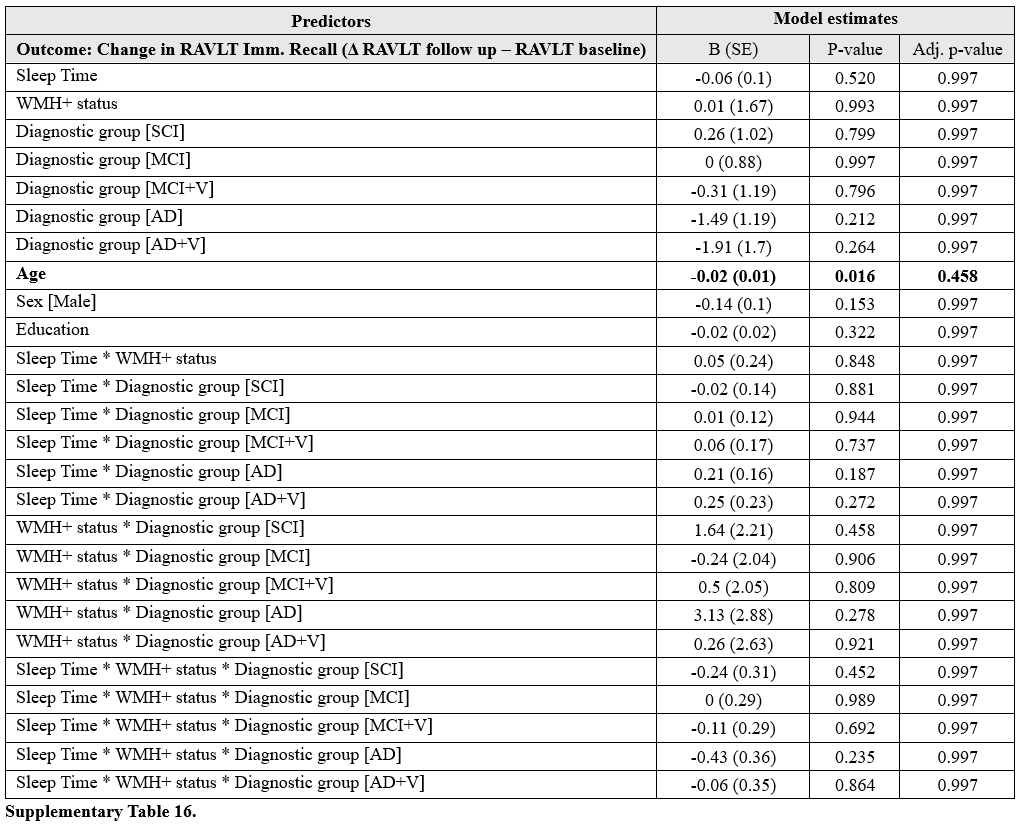

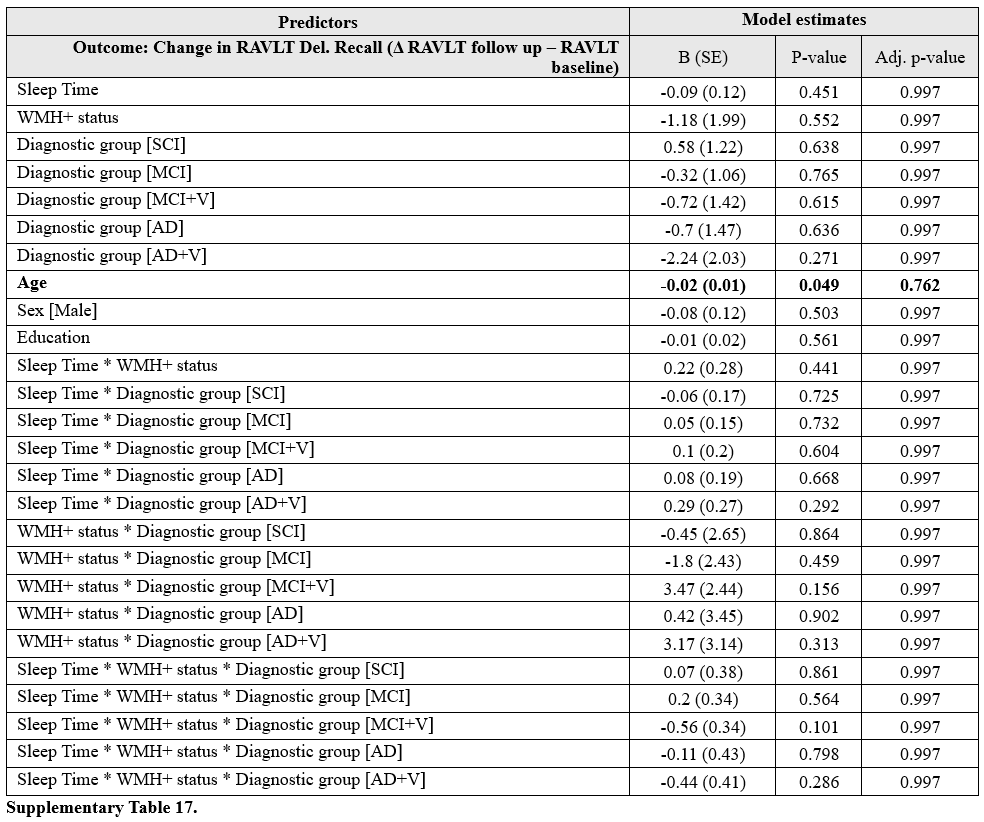
